# Povidone iodine, hydrogen peroxide and chlorhexidine mouthwashes reduce SARS-CoV2 burden in whole mouth fluid and respiratory droplets

**DOI:** 10.1101/2021.02.25.21252488

**Authors:** Bagavad Gita Jayaraman, Gunaseelan Rajan, Priya Kannian, Chandra Lavanya, Krittika Ravichandran, Nagalingeswaran Kumarasamy, Kannan Ranganathan, Veeraraghavan Aswini, Pasuvaraj Mahanathi, Stephen Challacombe, Jennifer Webster-Cyriaque, Newell W Johnson

## Abstract

SARS-CoV2 is transmitted primarily through oral mouth secretions and respiratory droplets. Commercial mouthwashes, povidone iodine (PI), hydrogen peroxide (HP) and chlorhexidine (CHX) have been tested in cell culture and RT-PCR studies for their efficacy to reduce SARS-CoV2 burden. Here, we evaluated SARS-CoV2 burden in whole mouth fluid (WMF) and respiratory droplets (RD) samples before and after the use of PI, HP or CHX mouthwashes in hospitalized COVID-19 patients using RT-PCR and rapid antigen test (RAT). Thirty-six SARS-CoV2 RT-PCR-positive in-patients were randomly assigned to one of the four groups: 20 and 60 minutes after 1% w/v PI or 1.5% HP; 90 and 180 minutes after 1.5% HP or 0.2% w/v CHX. WMF and RD samples were collected concurrently at baseline and after the two different time points. RD (92%) showed a higher reduction in SARS-CoV2 burden than WMF samples (50%; p=0.008). SARS-CoV2 burden was statistically lower at both 20 minutes (p=0.02) and 60 minutes (p=0.03) with PI; at 20 minutes with HP (p=0.0001); and 90 minutes with CHX (p=0.04). The overall and individual mean logarithmic reductions in the WMF and RD samples were greater than 1.0 at 20, 60 and 90 minutes after PI, HP or CHX. RAT-positive patients at 90 minutes post-treatment (n=3) demonstrated a one log increase in virus copies. Among the three RAT-negative post-treatment patients, SARS-CoV2 burden declined by one log in two while the third patient had a slight increase in RNA copies. In conclusion, we have shown for the first time that the mouthwashes, PI, HP and CHX can reduce the SARS-CoV2 burden in the concurrently collected RD and WMF samples. RAT is more appropriate than RT-PCR to evaluate the efficacy of the mouthwashes.

## Introduction

Whole mouth fluid (WMF) and respiratory droplets (RD) primarily transmit SARS-CoV2. *In vitro* cell culture and RT-PCR studies in WMF have shown that commercial mouthwashes - chlorhexidine (CHX), povidone iodine (PI) and hydrogen peroxide (HP) have significant virucidal activity against SARS-CoV2.^1,2^ However, RT-PCR of RNA viruses will detect both live and dead viruses, and should be interpreted cautiously. We have previously used rapid antigen testing (RAT) to detect SARS-CoV2 proteins in WMF thereby depicting its infectious state.^4^ This study evaluates SARS-CoV2 burden in WMF and RD samples before and after the use of PI, HP or CHX mouthwashes in hospitalized COVID-19 patients using RT-PCR and RAT.

## Methods

The study was approved by VHS-Institutional Ethics Committee (VHS-IEC/60-2020). Thirty-six SARS-CoV2 RT-PCR-positive (nasopharyngeal swab [NPS] and WMF) in-patients were randomly assigned after written informed consent to one of the four groups – 20 and 60 minutes after 1% w/v PI or 1.5% HP; 90 and 180 minutes after 1.5% HP or 0.2% w/v CHX. Early morning unstimulated drooled WMF samples and RD exhaled onto Whatman No.1 filter paper discs contained within surgical face masks were collected at baseline and at the two different time points after the corresponding mouthwash. Quantitative SARS-CoV2 RT-PCR was done using the automated QIAAmp Viral RNA kit (QIAGEN, Germany), Lightcycler 96 (Roche, USA) and a validated standard curve. SARS-CoV2 antigen in the WMF was tested in a subset of six patients before and after HP by RAT (SD Biosensor, Korea).^4^ Analyses (mean and standard deviations [SD]) and statistics (t-test and Fisher exact) were done using Microsoft Excel, VassarStats and Social Science Statistics.

## Results

Among the 36 patients, 13 (36%) were RT-PCR positive in RD samples at baseline. A 50% or greater decrease in the SARS-CoV2 copies from the baseline was considered significant. The reduction was significantly higher in RD (92%; 12/13) than WMF samples (50%; 18/36; p=0.008), perhaps due to lower baseline burden in RD samples (Table 1). Compared to the baseline, the mean copy numbers were statistically lower at both 20 minutes (p=0.02) and 60 minutes (p=0.03) with PI; at 20 minutes with HP (p=0.0001); and 90 minutes with CHX (p=0.04). The overall and individual mean logarithmic reductions (MLR) in the WMF and RD samples were greater than 1.0 at 20, 60 and 90 minutes after PI, HP or CHX (Table 1).

**Table 1:** Mean logarithmic reduction in SARS-CoV2 copies in the whole mouth fluid (WMF) and respiratory droplet (RD) samples by RT-PCR after the mouthwash

| Mouthwash | Baseline<br>Log copies<br>Mean±SD | Mean logarithmic reduction in SARS-CoV2 copies |  |  |  |
| --- | --- | --- | --- | --- | --- |
|  |  | 20 minutes<br>n/N (%) | 60 minutes<br>n/N (%) | 90 minutes<br>n/N (%) | 180 minutes<br>n/N (%) |
|  |  | Mean±SD | Mean±SD | Mean±SD | Mean±SD |
| *Whole mouth fluid (WMF)* |  |  |  |  |  |
| PI | 4.47±1.35 | 5/10 (50)<br>1.78±1.08 | 5/10 (50)<br>1.26±0.87 | ND | ND |
| HP | 5.03±0.79 | 6/10 (60)<br>1.17±0.27 | 5/10 (50)<br>1.62±1.64 | 2/8 (25)<br>1.45±1.45 | 2/8 (25)<br>0.93±0.82 |
| CHX | 5.18±1.89 | ND | ND | 5/8 (63)<br>1.62±1.23 | 2/8 (25)<br>0.43±1.51 |
| Overall | 4.89±1.34 | 11/20 (55)<br>1.48±0.68 | 10/20 (50)<br>1.44±1.26 | 7/16 (44)<br>1.53±1.34 | 4/16 (25)<br>0.68±1.17 |
| *Respiratory droplet (RD)* |  |  |  |  |  |
| PI | 2.50±0.40 | 3/3 (100)<br>2.5±0.40 | 2/3 (67)<br>1.58±1.87 | ND | ND |
| HP | 3.70±1.83 | 3/3 (100)<br>3.36±3.66 | 2/3 (67)<br>2.45±2.75 | 2/3 (67)<br>1.90±1.58 | 2/3 (67)<br>3.03±0.03 |
| CHX | 1.76±0.68 | ND | ND | 4/4 (100)<br>1.15±0.82 | 3/4 (75)<br>0.59±1.66 |
| Overall | 2.65±0.97 | 6/6 (100)<br>2.93±2.03 | 4/6 (67)<br>2.02±2.31 | 6/7 (86)<br>1.53±1.20 | 5/7 (71)<br>1.81±0.85 |

n – number of patients with a 50% or greater reduction in SARS-CoV2 copies compared to the baseline; N – number of patients tested with that particular mouthwash; % - percentage; ND – not done; PI – Povidone Iodine; HP – Hydrogen Peroxide; CHX – Chlorhexidine

WMF samples from six patients were tested for SARS-CoV2 antigens by RAT at baseline and at either 20 minutes (n=3) or 90 minutes (n=3) after HP. Six baseline samples were RAT-positive. Patients who remained RAT-positive at 90 minutes post-treatment (n=3) demonstrated a one log increase in virus copies (Table 2). Among the three patients who became RAT-negative post-treatment, in two individuals the SARS-CoV2 burden declined by one log. However, in the third patient a slight increase in RNA copies was detected.

**Table 2:** Comparison between RAT and RT-PCR for the determination of the reduction of SARS-CoV2 burden in whole mouth fluid samples before and after the use of hydrogen peroxide (HP)

| Patient No. | Time<br>(minutes) | <u>RAT</u> |  | <u>RT-PCR</u> |  | Log<br>difference* |
| --- | --- | --- | --- | --- | --- | --- |
|  |  | Baseline | After HP | Baseline | After HP |  |
| 1 | 20 | Positive | Negative | $3.10 \times 10^3$ | $2.50 \times 10^2$ | -1.1 |
| 2 | 20 | Positive | Negative | $2.12 \times 10^5$ | $6.36 \times 10^5$ | +0.5 |
| 3 | 20 | Positive | Negative | $5.47 \times 10^4$ | $3.87 \times 10^3$ | -1.1 |
| 4 | 90 | Positive | Positive | $1.98 \times 10^3$ | $3.77 \times 10^5$ | +2.3 |
| 5 | 90 | Positive | Positive | $2.04 \times 10^7$ | $3.25 \times 10^8$ | +1.2 |
| 6 | 90 | Positive | Positive | $5.03 \times 10^6$ | $1.22 \times 10^9$ | +2.4 |

HP – Hydrogen peroxide; RAT – rapid antigen test; RT-PCR – reverse transcriptase-polymerase chain reaction
\*A minus value in the log difference indicates decrease and a plus value indicates increase.

## Discussion

WMF and RD are primary modes of SARS-CoV2 transmission. Studies are reporting the efficacy of mouthwashes in reducing SARS-CoV2 burden in WMF. In this study, we report for the first time, reduction of SARS-CoV2 burden in RD for 20-90 minutes after PI, HP or CHX in addition to the concurrently collected WMF. This is consistent with *in vitro* cell culture and RT-PCR studies that have shown reduction in SARS-CoV2 burden.^2,3,5,6^ We also demonstrated for the first time that RAT is superior to RT-PCR for determining the efficacy of interventions designed to decrease oral transmission. The unbiased detection of viral RNA by RT-PCR irrespective of active viral infection can be problematic. The detection of antigen using RAT or ELISA based technologies allows mitigation of non-infectious/non-viable RNA that may be detected by RT-PCR.

## Data Availability

Data is not available in the links.

